# Impact of key meteorological parameters on the spread of COVID-19 in Mumbai: Correlation and Regression Analysis

**DOI:** 10.1101/2022.02.22.22271376

**Authors:** Sushant Shetty, Amit Gawade, Subodh Deolekar, Vaishali Patil, Rohit Pandharkar, Uday Salunkhe

## Abstract

**Purpose:** To understand key meteorological parameters that influence the spread of COVID-19 in Mumbai, India (based on data from April 2020 – April 2021).

**Methods:** The meteorological parameters chosen were Temperature, Dew Temperature, Humidity, Pressure, Wind Speed. The underlying basic relationships between meteorological parameters and COVID-19 information for Mumbai was understood using Spearman’s rank correlation coefficients. After establishing basic relationships, Linear analysis and Generalized Additive Model’s (GAM) were used to figure out statistically significant weather parameters and model them to explain the best possible variance in the pandemic data.

**Results:** A model of temperature and windspeed could explain 17.3% and 8.3% of variance in Daily new cases and Daily recoveries respectively. As for deaths occurring due to the virus, a model comprising of only pressure best explains a variance of 17.3% in the data. Non-Linear modelling based on GAM confirms the findings of linear analysis and establishes certain non-linear relationships as well.

**Conclusion:** SARS-CoV-2 belongs to the class of Human Coronaviruses (HCoV) which show seasonality depending on weather conditions. The above article focuses on understanding the underlying relationships between SARS-CoV-2 and meteorological parameters that would help progress basic research and formulation of policies around the disease for each weather/season.

**Competing interest:** The authors declare that they have no known competing financial interests or personal relationships that could have appeared to influence the work reported in this paper.

## 1. Introduction

At the time of writing, the total count of COVID-19 cases in India have crossed 32.4 million with the total number of related deaths exceeding 434,000. Mumbai in particular has had 741,389 cases with close to 16,000 deaths accounting for roughly 2.3% of the total cases and 2.15% of overall deaths (Municipal Corporation of Greater Mumbai, India 2020). Given the population density of the city which is 25,357 people/sq.km (Government of Maharashtra, India 2021) these percentages are significant since Mumbai is the economic capital of the country and can have a substantial impact on the overall recovery. First cases of the SARS-CoV-2 virus were reported in India on 30th January 2020 marking the start of the first wave. The first wave peaked around mid-September 2020 with daily cases reaching the 90,000 mark and had receded by January 2021 to a daily 15,000 mark. The second wave began around March 2021 and was more devastating than the first wave with daily cases reaching 400,000. The third wave is predicted to peak around mid-October 2021 but experts suggest it to be less severe than the former ones. Figure 1a and 1b show the progression of COVID-19 pandemic in Mumbai during the time period from 29th April 2020 to 21st April 2021. The few spikes in the Daily recoveries graph in Figure 1a are due to corrections received from the official data source. The Active cases graph in Figure 1b shows the first wave happening around March 2020, continuing until January 2021 and soon being followed by the second wave in March 2021.

**Figure 1.**
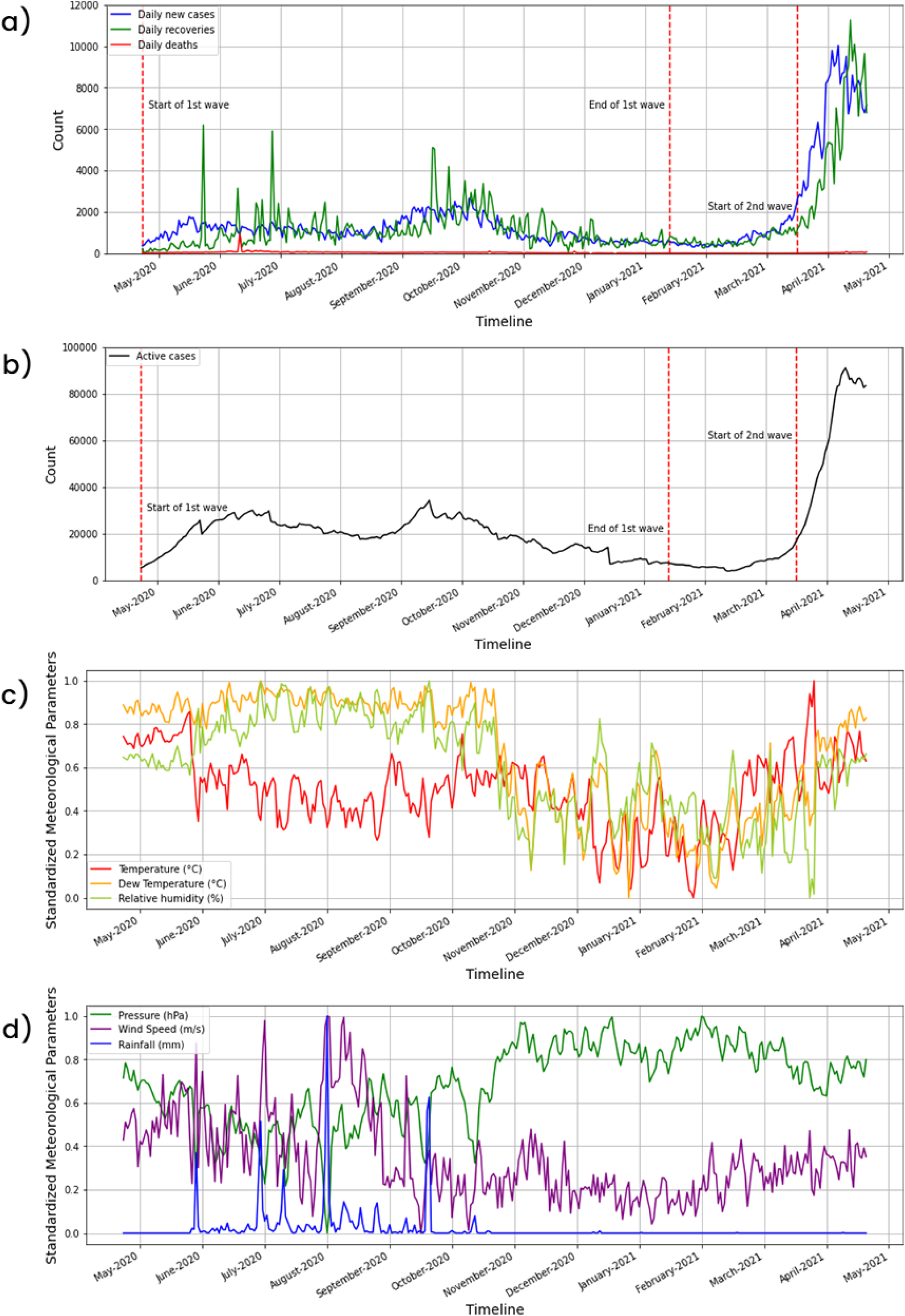
Time series data (29/04/2020 – 21/04/2021) of COVID-19 information and meteorological parameters: a) time series graph of Daily new cases, Daily recoveries and Daily deaths; b) time series graph of Daily deaths: c) standardized time series graph of Temperature, Dew Temperature and Relative Humidity; d) standardized time series graph of Pressure, Wind Speed and Rainfall

COVID-19 being a contagious disease, its spread can be attributed to multiple factors. The major factor contributing to the spread is human to human transmission via respiratory droplets in the atmosphere. A good metric to understand this factor is the R number. The R number in simple terms points to the number of people that an infected person can spread the disease to (Achaiah et al. 2020). So, a R number of 1.9 indicates that a single infected person can spread the disease to roughly 2 people. In January 2021, WHO put the number to be between 1.4 and 2.5. Other countries have reported much higher R numbers with China reporting a number between 4.7 - 6.6 (Gavi, The Vaccine Alliance 2020). However, R number is relative and depends on a lot of factors like total population, population density, immunity and variants. Other major factors contributing to the spread include rate of testing, quarantine periods, vaccination rates and lock down intensities among others. Associations between meteorological parameters and infectious diseases have been studied previously. Reports from such studies have shown high association between infection rates and weather parameters like temperature and relative humidity for influenza (Lowen et al. 2007). Common cold is reported to be seasonal in nature, with infection frequency increasing during autumn, peaking through to winter and then reducing in spring (Heikkinen 2003). Human coronaviruses (HCoV) namely 229E, NL63, OC42, HKU1 (Centers for Disease Control and Prevention 2020) were found to be seasonal indicating that they could be influenced by weather parameters (Monto et al. 2020). Given that SARS-CoV-2 belongs to a group of known coronaviruses, there exists a possibility that the spread of this virus peaks during favourable weather conditions. The objective of this study is to better understand the spread of COVID-19 under the influence of meteorological parameters in Mumbai, India.

The influence of meteorological parameters on infectious diseases has been studied extensively (Ma et al. 2010, Wang et al. 2020, du Prel et al. 2009, Zha et al. 2020, Tasci et al. 2018) and from these studies it has been established that weather parameters play an important role in assisting or resisting the spread of disease in a particular region. Table 1 [9 - 13] provides a summary of reports that have previously established relationships and strong correlations between diseases and weather parameters. Since human coronaviruses (HCoV) have been found to be seasonal (Park et al. 2020), there is a high possibility that SARS-CoV-2 virus which belongs to the same family might be affected by weather conditions in a similar way. Table 1 [15 - 23] provides a summary of recent studies conducted to understand the underlying relationships between weather parameters and the spread of COVID-19 virus in various countries.

**Table 1:** Summary of report on the influence of meteorological parameters on infectious diseases (1-5) and influence of meteorological parameters on COVID-19 spread (6 - 14)

|  |  | Area |  |  |
| --- | --- | --- | --- | --- |
| 1. | Is hand, foot and mouth disease associated with meteorological parameters? (Ma et al. 2007) | Hong Kong | Collinearity adjusted data showed positive correlation between HFMD consultation rates (with a 2-week lag time) and weather parameters namely temperature (mean), diurnal temperature difference, humidity and wind speed. As per sensitivity analysis, HFMD consultation rate was majorly influenced by humidity whereas wind speed had the least effect. | temperature (mean), diurnal temperature difference, humidity |
| 2. | The long-term effects of meteorological parameters on pertussis infections in Chongqing, China, 2004–2018. (Wang et al. 2020) | Chongqing, China | Statistical analysis of data showed that pertussis is a seasonal disease which spikes during spring and summer. Major weather parameters affecting the spread of the disease are temperature (1°C increase causes 19.496% increase in cases), precipitation (10 mm increase causes 3.641% increase in cases), air pressure (1hPa decrease causes 3.559% decrease in cases) and wind velocity (1 m/s increase causes 3.812% increase in cases). | Temperature, pressure |
| 3. | Are Meteorological Parameters Associated with Acute Respiratory Tract Infections? (du Prel et al. 2009) | Mainz, Germany | Infectious pathogens like influenza virus and respiratory syncytial virus (RSV) show annual/biannual rhythmicity mainly during the cold season. RSV, influenza A, rhinovirus and adenovirus were negatively correlated with temperature. On the other hand, rhinovirus showed positive correlation with humidity. | Temperature, humidity, wind speed |
| 4. | Effects of meteorological factors on the incidence of mumps and models for prediction, China. (Zha et al. 2020) | China | Rainfall, pressure, average temperature and wind speed showed major influence on the occurrence of mumps in China. Factors like temperature, humidity and wind speed were positively correlated whereas air pressure showed negative correlation. | Temperature, humidity, wind speed, pressure |
| 5. | Relationship of Meteorological and Air Pollution Parameters with Pneumonia in Elderly Patients. (Tasci et al. 2018) | Turkey | Temperature was negatively correlated whereas humidity, rainfall and sea-level pressure were positively correlation with admittance of pneumonia patients. | Temperature, humidity, rainfall and sea-level pressure |
| 6. | A systematic review and meta-analysis on correlation of weather with COVID-19. (Majumder and Ray 2021) | 110 countries | Temperature, humidity, wind speed and rainfall showed positive correlation whereas UV radiation showed positive correlation with COVID-19 infection rate. In terms of deaths due to COVID-19 humidity showed higher significance and negative correlation. | Temperature, Humidity, Wind Speed and Rainfall |
| 7. | Association between meteorological factors and daily new cases of COVID-19 in 188 countries: A time series analysis. (Yuan et al. 2021) | 188 countries | actors like mean temperature, wind speed and relative humidity show negative correlation whereas diurnal temperature range showed positively correlated with daily new cases of COVID-19. The associations were further evident with temperature and relative humidity in the lower ranges (21.07°C and 66.83% respectively). | Temperature (mean and diurnal), Wind Speed, Relative Humidity |
| 8. | Impact of meteorological factors on COVID-19 pandemic: Evidence from top 20 countries with confirmed cases. (Sarkodie and Owusu 2020) | 20 countries | Temperature and relative humidity show negative correlation with incidence of COVID-19 cases. Wind speed, surface pressure, dew point and precipitation were observed to have a positive correlation with COVID-19 incidence and deaths, and had a negative correlation with recovery rates. | Temperature, Relative Humidity, Wind Speed, Surface Pressure, Dew point, Precipitation |
| 9. | Meteorological parameters and COVID-19 spread-Russia a case study. (Shankar et al. | Russia | Temperature, dew point and humidity among other weather parameters showed major influence on COVID-19 conditions. Humidity was observed to have negative correlation | Temperature, Dew Point, Humidity |
|  | 2021) |  | whereas temperature and dew point showed positive correlation with death rates. Humidity also showed negative association with daily new COVID-19 cases. |  |
| 10. | A correlation study between meteorological parameters and COVID-19 pandemic in Mumbai, India. (Kumar and Kumar 2020) | Mumbai, India | Factors like temperature (Tmin), dew point (DPmax), relative humidity and surface pressure show more statistical significance compared to other meteorological parameters. Among these, relative humidity and pressure showed more influence on the active number of COVID-19 cases. | Temperature, Dew Point, Relative Humidity and Surface Pressure |
| 11. | Association of air pollution and meteorological variables with COVID-19 incidence: Evidence from five megacities in India. (Kolluru et al. 2020) | Bangalore, Chennai, Delhi, Kolkata and Mumbai: Indi | Among weather parameters, temperature showed major correlation with COVID-19 cases ad deaths. The parameter could also explain up to 34% and 30% variability in confirmed cases and deaths. Other significant parameters were wind speed and relative humidity which explained 11% and 4% variance in COVID-19 cases respectively. | Temperature, Wind Speed and Relative Humidity |
| 12. | Impact of meteorological parameters on COVID-19 transmission in Bangladesh: a spatiotemporal approach. (Hridoy et al. 2021) | Bangladesh | The study uses relative risks to gauge the influence of meteorological parameters on COVID-19 incidence. RR of mean temperature, wind speed and humidity were calculated to be 1.222[95% CI(1.214- 1.232)], 1.087[95% CI(1.083-1.090)] and 1.027[95% CI(1.025-1.029)] respectively | Temperature (mean), Wind Speed and Humidity |
| 13. | Impact of Meteorological Conditions on the Dynamics of the COVID-19 Pandemic in Poland. (Bochenek et ak. 2021) | Poland | Factors such as maximum temperature, sunshine duration, relative humidity and variance in mean daily temperature showed higher statistical significance on COVID-19 spread. Temperature, sunshine hours and humidity (with a 14-day lag) showed positive correlation with confirmed cases. As for deaths due to COVID-19, humidity showed negative correlation whereas sunshine hours were observed to have a positive correlation. | Temperature (max), Sunshine duration and Relative Humidity |
| 14. | Association of COVID-19 pandemic with meteorological parameters over Singapore (Pani et al. 2020) | Singapore | Out of the 9 used meteorological parameters namely temperature, dew point, air pressure, relative and absolute humidity, water vapor, wind speed, boundary layer height and ventilation coefficient, COVID-19 spread was positively correlated with temperature, dew point, relative and absolute humidity, and water vapor. | Temperature, Dew Point, Humidity (relative and absolute) |

## 2. Material and Methods

This retrospective study examining the impact of meteorological parameters on the spread of SARS-CoV2 virus was done on data collected from 29th April 2020 to 21st April 2021 for Mumbai, India. Although COVID-19 cases were detected in the country way before the starting date of the study, reliable and continuous data for Mumbai was available on the official website from 29th April, 2020 and hence was chosen as the starting point. COVID-19 data for Mumbai was obtained from the city’s municipal corporation (Municipal Corporation of Greater Mumbai, India 2020) and state government’s (Government of Maharashtra, India 2022) dashboard. The dashboards contain 4 standard datapoints useful for this study, namely Daily new cases, Daily recoveries, Daily deaths and Active cases. The city’s municipal corporation is divided into different wards which update this information on a daily basis. This information is then collated and released by the municipal corporation on a daily dashboard. The data has been expressed graphically in Fig 1a and 1b.

The meteorological data for the aforementioned time period was obtained using OpenWeatherMap’s API (OpenWeatherMap 2021). These included parameters like temperature, relative humidity, pressure, wind speed and rainfall on an hourly basis. Hourly rainfall values were added whereas the rest of the parameters were averaged to get the daily values. Dew temperature for the region was calculated from the daily values using the simple formula:

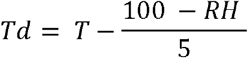

where:

**Td** = Dew temperature

**T** = Average temperature

**RH** = Relative humidity

Being a coastal city that lies in the northern hemisphere closer to the equator, Mumbai has high relative humidity and high average seasonal temperatures. It experiences summer from February to May, a rainy season from June to September and winters from October to January. Fig 1c and 1d graphically represent the standardized weather parameters for Mumbai for the study time period. The values of all the weather parameter were standardized based on their individual mean and standard deviation values for better graphical representation.

The basic association between COVID-19 information and meteorological parameters was examined using Spearman’s rank correlation coefficient since it can detect monotonic patterns in the data unlike Pearson’s correlation. Pearson’s coefficient is used to quantify the linear relationship between two variables and is calculated as follows:

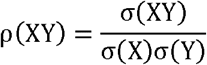

where:

σ(XY) = covariance of X and Y

σ(X) = standard deviation of X

σ(Y) = standard deviation of Y

Spearman’s rank correlation coefficient on the other hand ranks the values of two variables in ascending order and calculates the Pearson coefficient of the rank values. Due to the use of ranks to calculate correlation, Spearman’s method is able to identify monotonic relations in the data as opposed to just linear relations which the Pearson’s method can identify.

Since weather is a highly complex interdependent phenomenon, weather parameters are bound to be correlated with each other meaning that meteorological data can be highly collinear. Collinearity among independent variables may lead to incorrect conclusions when establishing relationships with dependent variables (Næs and Mevik 2001). To tackle the issue of collinearity, parameters were grouped together such that they had the least correlation amongst themselves. These parameters were then entered in a step wise manner to find the parameters with the best fit for the data. Theoretically speaking, reported infections lag behind the actual infection day due to the incubation period of the virus and the delay in seeking medical help. Reported incubation period for coronavirus typically ranges from 1-14 days with the median incubation period in India being 5.1 days (Government of India 2021). To account for this delay between actual infection, incubation period and time taken to seek medical assistance, a lag of 5-14 days was introduced in the data for daily new cases to ascertain the relationship. The regression analysis was done using SPSS (version 25.0) and a p-value of less than 0.05 was chosen to establish statistical significance. In the second part of the analysis, to account for non-linear relationships between the weather parameters and COVID-19 data, Generalized Additive Models (GAM) were used. GAM is an extension of linear modelling wherein it allows linear models to learn nonlinear relationships present in the data. Instead of using parameters weights to sum up a linear model, GAM’s use sum of arbitrary functions to model a variable. It does so by using a connection function (Yuan 2021) that fits the data in a wide variety of shapes and sums them up to get the best fit.

## 3. Results

Correlation coefficients can be interpreted as follows:

1. – 0.10 (Negligible correlation)
2. 0.10 – 0.39 (Weak correlation)
3. 0.40 – 0.69 (Moderate correlation)
4. 0.70 – 0.89 (Strong correlation)
5. 0.9 – 1.00 (Very strong correlation)

The Spearman’s rank correlation coefficient between individual COVID-19 data columns and meteorological parameters are shown below:

1. Daily new cases: Temperature (0.49), Dew Temperature (0.48), Pressure (−0.51), Humidity (0.37), Windspeed (0.23), Rainfall (0.21)
2. Daily recoveries: Temperature (0.17), Dew Temperature (0.3), Pressure (−0.29), Humidity (0.28), Windspeed (−0.016), Rainfall (0.17)
3. Daily deaths: Temperature (0.23), Dew Temperature (0.81), Pressure (−0.77), Humidity (0.76), Windspeed (0.41), Rainfall (0.62)

Daily new cases, Daily deaths and Active cases show positive correlation with temperature, dew temperature, humidity, wind speed and rainfall, and are negatively correlated with pressure. Daily recoveries show positive association with temperature, dew temperature, humidity and rainfall, and negative association with pressure and wind speed. From this it can be inferred that most meteorological parameters have moderate to strong associations with the spread of COVID-19 in the city. Among all the weather parameters, pressure is negatively correlated with all COVID-19 variables. Rest of the variables are all positively correlated with COVID-19 information. This preliminary analysis using correlation coefficients shows the underlying relationship between meteorological parameters and COVID-19 information. These relationships are further explored in the upcoming sections using Regression Analysis and Generalized Additive Modelling. Spearman’s rank correlation coefficient amongst meteorological parameters shows high levels of collinearity among pressure, dew temperature, humidity and rainfall (negative correlation greater than 0.7). Hence, among the above-mentioned parameters, only pressure was considered for regression analysis. Apart from pressure, temperature and wind speed were also included for regression analysis.

Regression Analysis was done individually on Daily new cases, Daily recoveries and Daily deaths. Due to the problem of high collinearity amongst variables, only temperature, pressure and wind speed were considered for the analysis. Knowing the relationship between pressure and COVID-19 variables is enough to establish relationships between dew temperature, humidity, rainfall and COVID-19 data. Table 2 shows regression analysis done for Daily new cases (with multiple time lags), Daily recoveries and Daily deaths.

**Table 2:** Summary of Linear Analysis and Generative Additive Models of meteorological parameters vs COVID-19 information.

| Regression analysis |  |  |  |  |  |  |
| --- | --- | --- | --- | --- | --- | --- |
| COVID-19 Parameter | Model | Parameters included | Partial correlation coefficient | p-value of parameters | R<sup>2</sup> (variance) for the model | p-value for the model |
| Daily new cases | M0 | Temperature | 0.365 | 0.00 | 0.133 | 0.00 |
|  | M1(7 days) | Temperature | 0.379 | 0.00 | 0.144 | 0.00 |
|  | M2 (10 days) | Temperature | 0.416 | 0.00 | 0.173 | 0.00 |
|  |  | Windspeed | -0.111 | 0.036 |  | 0.000 |
|  | M3 (14 days) | Temperature | 0.37 | 0.000 | 0.137 | 0.000 |
| Daily recoveries |  | Temperature | 0.275 | 0.000 | 0.086 | 0.000 |
|  |  | Windspeed | -0.036 | 0.036 |  |  |
| Daily deaths |  | Pressure | -0.416 | 0.000 | 0.173 | 0.000 |
| **Generalized Additive Models (GAM)** |  |  |  |  |  |  |
| Daily new cases |  | Temperature |  | 0.00 | 0.368 |  |
|  |  | Pressure |  | 0.03 |  |  |
|  |  | Windspeed |  | 0.00 |  |  |
| Daily recoveries |  | Temperature |  | 0.00 | 0.163 |  |
|  |  | Pressure |  | 0.07 |  |  |
|  |  | Windspeed |  | 0.011 |  |  |
| Daily deaths |  | Pressure |  | 0.000 | 0.184 |  |

1. Daily new cases: M0 which refers to analysis done with no lag period shows positive correlation with temperature, has a p-value of 0.000 and explains 13.3% of the variance (indicated by R^2^) in new cases. Similarly multiple models were built with different lag periods: M1(7 days), M2(10 days), M3(14 days). Incubation period for SARS-CoV-2 virus is 5.1 days and assuming that infected people take 1-3 days to seek medical help and 1 more day to get the test reports, the ideal lag time would be ∼10 days. In line with this, the partial correlation coefficients for M2 are higher than the rest of the models and it also explains the variance(R^2^) in the data much better (17.3%) than the other models. Hence model M2 was chosen as the best fitting model to explain the trends in Daily new cases. From the partial correlation coefficients for M2, it is observed that Daily new cases have a moderate positive (0.416) association with temperature and weak negative (−0.111) association with wind speed. The black lines in Figure 2a and 2b show the linear relation expressed by the partial correlation coefficients.
2. Daily recoveries: Regression analysis for recoveries shows temperature and windspeed as the key influential parameters. A combined model of temperature and wind speed best explains the variance (8.7%) in daily recoveries. Figure 6a and 6b indicate this association with black lines where Temperature is positively correlated (0.275) and Wind Speed is negatively correlated (−0.036) with Daily recoveries.
3. Daily deaths: For Daily deaths, only pressure has a negative association (−0.416) and explains 17.3% variance happening over the period of study. The moderate association of pressure with daily deaths is shown in Figure 2e as a black line with an apparent downward trend.

**Figure 2.**
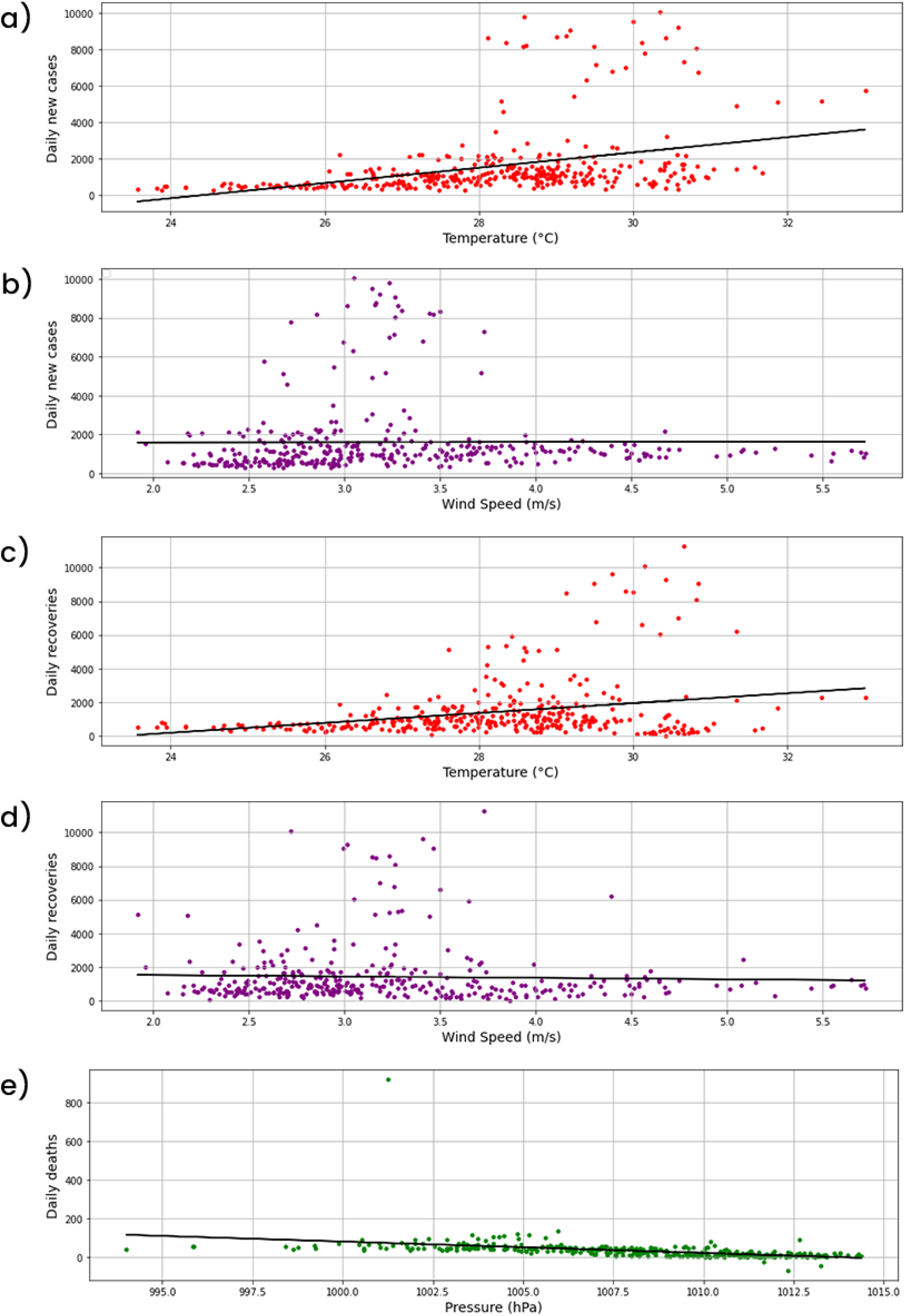
Linear analysis results of meteorological parameters vs COVID-19 information: a) Temperature vs Daily new cases (correlation coefficient = 0.416); b) Wind Speed vs Daily new cases (correlation coefficient = -0.111); c) Temperature vs Daily recoveries (correlation coefficient = 0.275); d) Wind Speed vs Daily recoveries (correlation coefficient = -0.036); e) Pressure vs Daily deaths (correlation coefficient = -0.416)

Multivariate analysis using GAM was conducted using the same three meteorological parameters since they expressed most of the variance in the meteorological data. On the x-axis are the meteorological parameter values while the vertical axis contains the contribution of the connection function to the shaped values.

1. Daily new cases: Using GAM, it can be seen from Figure 3a, 3b and 3c that temperature has a positive correlation with Daily new cases beyond 30°C whereas for pressure there is slight negative correlation beyond roughly 1009 hPa. Wind speed shows slight positive correlation up to 3.2 m/s beyond which the relation become non-linear. A combined model of these parameters can explain 36.8% of variance in Daily new cases indicated by R^2^ of the overall model in Table 2.
2. Daily recoveries: The GAM summary for Daily recoveries shows that a model comprising of temperature, pressure and wind speed can explain 16.3% of the variance in the data. The graphs in Figure 3d, 3e and 3f show the relationships between the parameters and Daily recoveries. Temperature shows a strong positive correlation with Daily recoveries for the entire value range. Pressure shows non-linear trend up until 1010bhPa beyond which it shows a downward trend. Wind speed graph shows no particular trend in the data and can be considered to be non-linear.
3. Daily deaths: This feature is negatively correlated with pressure beyond roughly 1000 hPA as can be seen in Figure 3g. This model can explain 18.4% of the variance in the data as can be seen by the value of R^2^ in Table 2.

**Figure 3.**
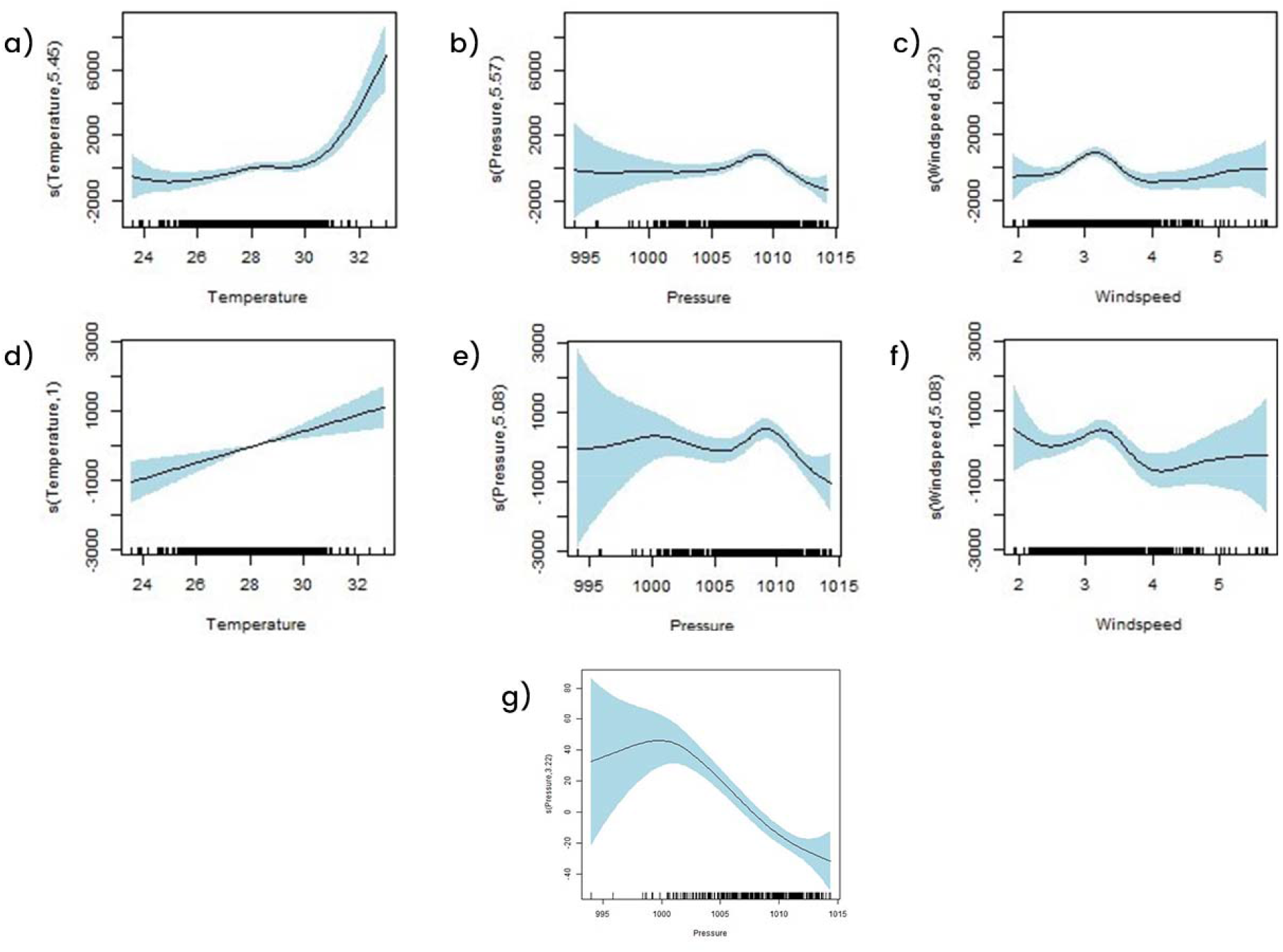
Generalized Additive Modes for COVID-19 information vs meteorological parameters: a) Temperature graph for Daily new cases; b) Pressure graph for Daily new cases; c) Wind Speed graph for Daily new cases; d) Temperature graph for Daily recoveries; e) Pressure graph for Daily recoveries; f) Wind Speed graph for Daily recoveries; g) Pressure graph for Daily deaths

## 4. Discussion

Although SARS-CoV-2 virus has infected most of the countries in the world, its incidence and intensity vary with time and place. Hence studies conducted in different countries could differ from each other on account of the different weather conditions and administrative actions taken to contain the spread. Another factor which might lead to different interpretations in results is the contrast in incubation period and R number reported in different countries. A study (Tushabe 2020) suggests that the virus could be more effective (in terms of infection rates and deaths) in countries that are located in the temperate region. However, this study was conducted during the initial days of the pandemic and could lead to erroneous conclusions. Also, it is difficult to ascertain the findings from this study in India, since the country lies in the temperate as well as the tropical region. Although an extensive and timely study would be required to establish seasonality, preliminary studies in this report suggest a certain rhythmicity in infection rates with cases peaking during the summer season (March - September), plateauing during the winter season (October to February) and again rising during the next summer season.

Multiple studies have tried to explain the association between the spread of contagious diseases and meteorological parameters (Ma et al. 2010, Wang et al. 2020, du Prel et al. 2009, Zha et al. 2020, Tasci et al. 2018). Most of these studies have established a strong association between weather related parameters and the disease. And since SARS-CoV-2 virus belongs to a group of known Human Coronaviruses (HCoV) which are seasonal, there are high chances that the weather in a particular region can influence the occurrence of the COVID-19 infection. However, since this is a new virus with effects and consequences different from its family, more extensive research needs to be done to establish facts about the influence of weather on COVID-19 spread. Various studies on components affecting the spread of the pandemic in India were also conducted (Majumder and Ray 2021, Yuan et al. 2021, Sarkodie and Owusu 2020, Shankar et al. 2021, Kumar and Kumar 2020, Kolluru et al. 2020, Hridoy et al. 2021, Bochenek et al. 2021, Pani et al. 2020). Preliminary studies done in this matter indicate some level of influence in cities like Delhi and Indore where cases showed moderate associations with temperature levels (Roy 2021). In case of Pune, relative humidity was found to have a positive association with daily infections with a lag period of one week (Roy 2020). COVID-19 incidence in cities like Bengaluru and Kolkata showed high correlation with temperature, and parameters like wind speed and relative humidity could explain certain levels of variance in the data (Kolluru et al. 2020). On the basis of Spearman’s correlation method, a study conducted in Mumbai highlighted relative humidity and pressure as the key factors influencing the number of active cases (Kumar and Kumar 2020). Since Mumbai lies in the tropical region with average temperatures usually being on the higher side, the conclusions from this study suggest that Mumbai might be more susceptible to infections since cases tend to rise with a rise in the temperature.

The above study observed high statistical correlation between certain meteorological parameters and the spread of COVID-19 in Mumbai, India. Temperature, Wind Speed and Pressure in particular were observed to be the key factors explaining the change in COVID-19 data on a daily basis. Although it should be noted that the information conveyed by these parameters in terms of variance differed for linear analysis and GAM’s, and was usually on the lower side. This could be attributed to the fact that infectious diseases are influenced by other factors such as population immunity, variants of the pathogen and measures taken to curb the spread e.g., lock downs and vaccination drives. However, the variance amount explained by meteorological parameters points out to the fact that like other infectious diseases, SARS-CoV-2 virus too is influenced by the climate and that weather parameters should be taken into consideration while understanding its future course.

## Data Availability

All data produced in the present study are available upon reasonable request to the authors

---

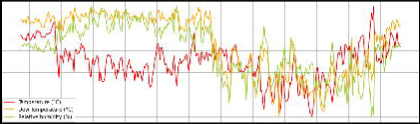

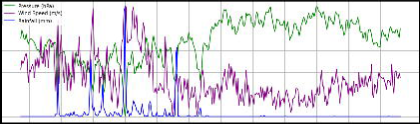

## Notes

### Competing Interest Statement

The authors have declared no competing interest.

### Funding Statement

This study did not receive any funding

### Author Declarations

Only open source data available from Mumbai city's municipal corporation website and Maharashtra state government website were used for this study. Links: 1.https://stopcoronavirus.mcgm.gov.in/assets/docs/Dashboard.pdf 2.https://www.covid19maharashtragov.in/mh-covid/dashboard

